# Interrogating DNA methylation associated with Lewy body pathology in a cross brain-region and multi-cohort study

**DOI:** 10.1101/2025.03.13.25323837

**Authors:** Joshua Harvey, Jennifer Imm, Morteza Kouhsar, Adam R. Smith, Byron Creese, Rebecca G. Smith, Gregory Wheildon, Valentin Laroche, Szi Kay Leung, Leonidas Chouliaras, Gemma Shireby, Zane Jaunmuktane, Eduardo De Pablo-Fernández, Thomas Warner, Debra J. Lett, Djordje Gveric, Hannah Brooks, Johannes Attems, Alan Thomas, Wilma DJ Van de Berg, Emma Dempster, Clive Ballard, John T O’Brien, Dag Aarsland, Jonathan Mill, Lasse Pihlstrøm, Ehsan Pishva, Katie Lunnon

## Abstract

Lewy body (LB) diseases are an umbrella term encompassing a range of neurodegenerative conditions all characterized by the hallmark of intra-neuronal α-synuclein associated with the development of motor and cognitive dysfunction. In this study, we have conducted a large meta-analysis of DNA methylation across multiple cortical brain regions, in relation to increasing burden of LB pathology. Utilizing a combined dataset of 1,217 samples across 844 unique donors, we identified a set of 24 false discovery rate (FDR) significant loci that are differentially methylated in association with LB pathology, the most significant of which were located in *UBASH3B* and *PTAFR*, as well as an intergenic locus. Ontological enrichment analysis of our meta-analysis results highlights several neurologically relevant traits, including synaptic alterations. We leverage our data to compare DNA methylation signatures between different neurodegenerative pathologies and highlight a shared epigenetic profile across LB diseases, Alzheimer’s disease and Huntington’s disease, although the top-ranked loci show disease specificity. Utilizing summary statistics from previous large-scale genome-wide association studies we identified significant enrichment of DNA methylation differences with respect to increasing LB pathology in the *SNCA* genomic region. We identified specific relationships between genetic risk variants for LB-Dementia and Parkinson’s disease to methylation quantitative trait loci in the promoter region and expression of SNCA transcript isoforms.

## INTRODUCTION

The Lewy body diseases including Parkinson’s disease (PD) and Dementia with Lewy bodies (DLB), are neurodegenerative diseases classified by the accumulation of alpha-synuclein (α-synuclein) in neurons, forming Lewy bodies (LBs). Collectively these diseases are the second most common cause of neurodegeneration and dementia, following Alzheimer’s disease (AD) and confer a high care giver burden, given the complex nature of the disease and symptoms^1^. According to diagnostic criteria, DLB and Parkinson’s disease dementia (PDD) are both defined as Lewy Body Dementia (LBD), but are distinguished clinically based on the timing of onset of parkinsonism and dementia symptoms^2^. At post-mortem examination these disorders present similar underlying pathological profiles, with, in addition to LBs, mixed pathology of neurofibrillary tangles (NFT) and amyloid beta (Aβ) plaques commonly featuring^3^.

Over the last few years, more studies have been undertaken to investigate the genetic mechanisms underpinning PD^4^ and LBD^5^. These studies have highlighted numerous risk loci associated with these diseases, including 78 loci in PD^4^ and five genome wide significant loci in LBD, three being known PD risk genes (*SNCA*, *GBA* and *TMEM175*) and two being AD risk genes (*APOE* and *BIN1*)^5^. However, whilst these studies have provided novel insights into disease pathways, genetic variation only explains a small amount of disease susceptibility, with environmental factors also being robustly associated with disease development and progression^6–8^. In recent years, there has been a growing appreciation that epigenetic mechanisms may mediate the interaction between genetic and environmental risk factors in the etiology of sporadic neurodegenerative diseases. Indeed, several epigenome-wide association studies (EWAS) have now been performed in AD, which have culminated in several large-scale meta-analyses, nominating numerous robust and reproducible alterations in DNA methylation in the cortex during disease^9–12^. However, there has been considerably less research exploring DNA methylomic signatures in relation to LB disease, with in fact only one EWAS performed to date with a substantial sample size (n > 100) and utilizing multiple independent cohorts^13^. That study utilized the Illumina Infinium MethylationEPIC array to quantify DNA methylation in the prefrontal cortex (PFC) in a primary discovery analysis from 73 control and 249 LB disease spectrum patients from the Netherlands Brain Bank (NBB). Their analysis focused on changes associated with LB pathology development, irrespective of the primary diagnosis and reported 24 loci significantly associated with Braak LB stage, with a strong concordance of the direction of effect with findings from the independent Brains for Dementia Research (BDR) cohort. Among the reported loci, four CpG sites were significant in both cohorts, annotated to *TMCC2*, *SFMBT2*, *AKAP6*, and *PHYHIP*.

To investigate the epigenetic underpinnings of these diseases in relation to neuropathology, we have designed the largest EWAS of LB disease to date, which includes multiple cortical brain regions and a meta-analysis with three independent cohorts. Our analyses utilized 1,217 samples from 844 unique donors and identified 24 false discovery rate (FDR) significant loci associated with LB pathology in the cortex. Leveraging prior studies of DNA methylation undertaken in AD and Huntington’s Disease (HD), we found shared epigenomic profiles between neurodegenerative diseases. Finally, we identified sites with evidence of genetic influence on DNA methylation levels and demonstrate an enrichment of methylation in the *SNCA* gene in LB disease. We observed a strong colocalization of genetic risk loci in the 5’ region of the gene, captured in both LBD and PD GWAS, to cortical methylation quantitative trait loci (mQTLs) for multiple methylation loci.

## RESULTS

### Meta analysis of differential DNA methylation associated with cortical Lewy bodies

Utilizing a dataset comprising three independent cohorts with DNA methylation profiling conducted on the Illumina Infinium MethylationEPIC v1.0 array, we sought to test for associations with the distribution of LB pathology, as measured by the Braak LB staging criteria. The cohort included novel data generated from the UK Brain Bank Network (UKBBN, n = 779 samples, 406 unique donors, n = 387 Anterior Cingulate Cortex (ACC), n = 392 Prefrontal Cortex (PFC)) and data from two existing datasets: the Netherlands Brain Bank (NBB, n = 322 PFC donors)^13^ and Brains for Dementia Research (BDR, n = 116 PFC donors)^14^ (**Figure 1**). Separately for each cohort, we used linear regression to identify differentially methylated positions (DMPs) associated with LB pathology, including confounding variables (age, sex, cell type proportions, technical batch variables, post-mortem interval and Braak NFT stage) and surrogate variables as required to ensure that the genomic inflation lambda value < 1.2 (**Methods**, **Supplementary Figure 1**). All donors in this dataset were either controls with a pathological confirmation of lacking neurodegenerative disease or required a primary neuropathologically confirmed diagnosis of incidental LB disease, PD, PDD or DLB (**Methods**). Given that 50-80% of LB disease have concomitant AD pathology^15^ where possible we minimized the number of individuals with advanced Braak NFT stage (> III) in our cohorts based on inclusion criteria for a neuropathological primary diagnosis of LB disease (**Figure 1, Supplementary Table 1, Methods**). We further included Braak NFT as a covariate in our primary discovery models (**Methods**). We conducted a fixed effect, inverse variance weighted meta-analysis of the three cohorts (total sample size = 1,217, unique donors = 844) for 774,310 methylation sites passing data quality control (QC). We identified three DMPs associated with LB pathology stage at a genome-wide significance association threshold of p < 9 x 10^-8^ (**Figure 2A&B**), whilst a further 21 DMPs were identified at a more lenient false FDR corrected p-value threshold (p_adj_ < 0.05) (**Figure 2A, Supplementary Table 2**). The most significant locus was annotated to probe cg13847853, which is a DMP with previous significant association to AD NFT pathology^12,13^ and is novel in the context of LB disease. It represents an intergenic locus located between the protein coding genes *KRT19* (-11,777 base-pairs) and *KRT9* (+31,973 base-pairs). It is also proximal to the long non-coding RNA gene *LINC00974* (+ 9,522 base-pairs).

**Figure 1.**
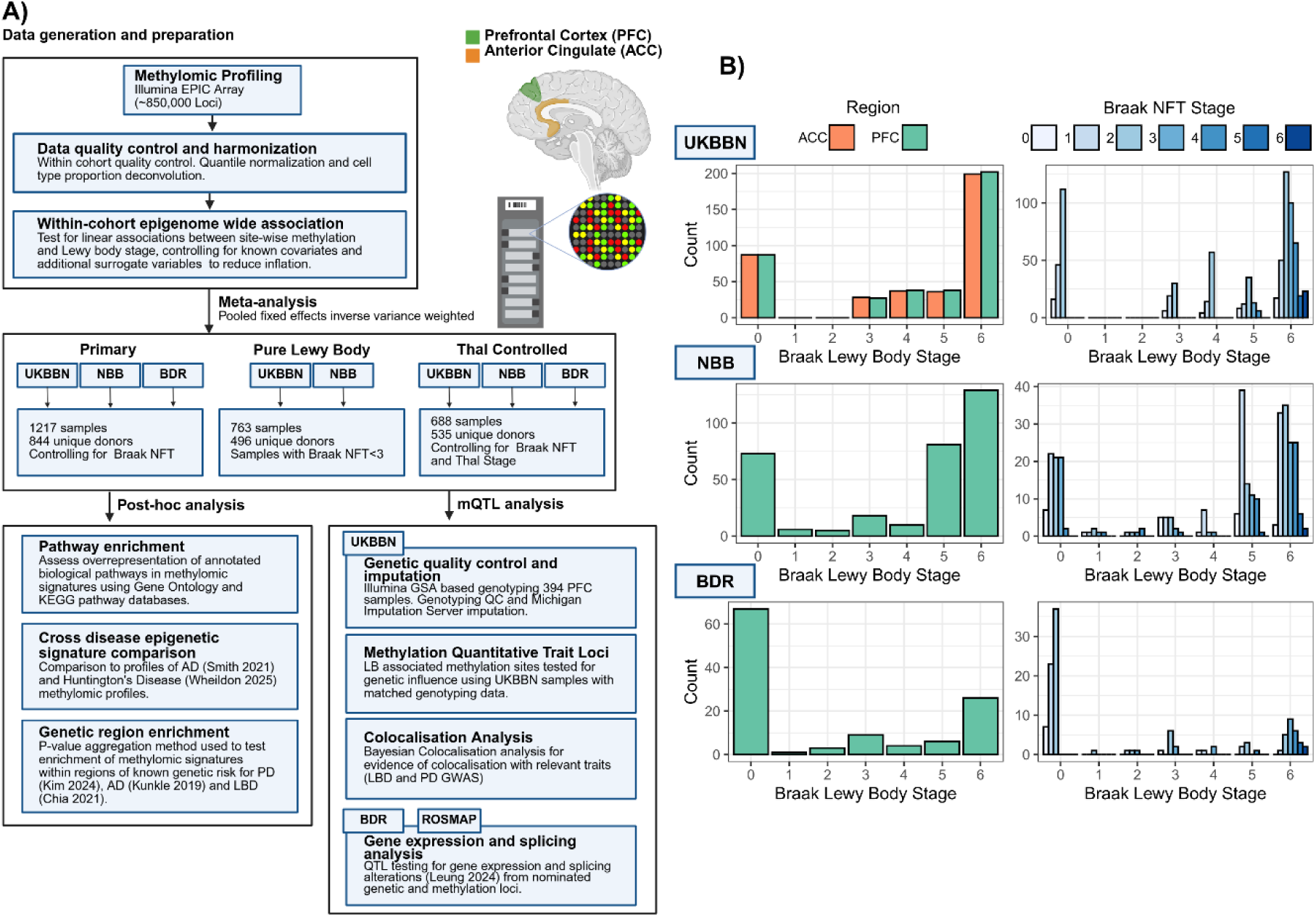
Summary of cohorts and samples used in DNA methylation association meta-analysis of Lewy bodies. **A)** Brain regions and analysis pipeline for meta-analyses. The brain regions include two cortical regions, the prefrontal cortex (PFC, BA9) and the anterior cingulate cortex (ACC, BA24). The analysis plan outlining steps in data generation, harmonization, within cohort linear association analyses and fixed effects meta-analysis. **B)** Summary of sample numbers in the UK Brain Bank Network (UKBBN), Netherlands Brain Bank (NBB) and Brains for Dementia Research (BDR) cohorts. Left hand bar plots show number of samples per Braak LB stage, right hand plots show number of samples with coinciding AD pathology as measured by the Braak NFT stage. Components of figure created in https://BioRender.com.

**Figure 2.**
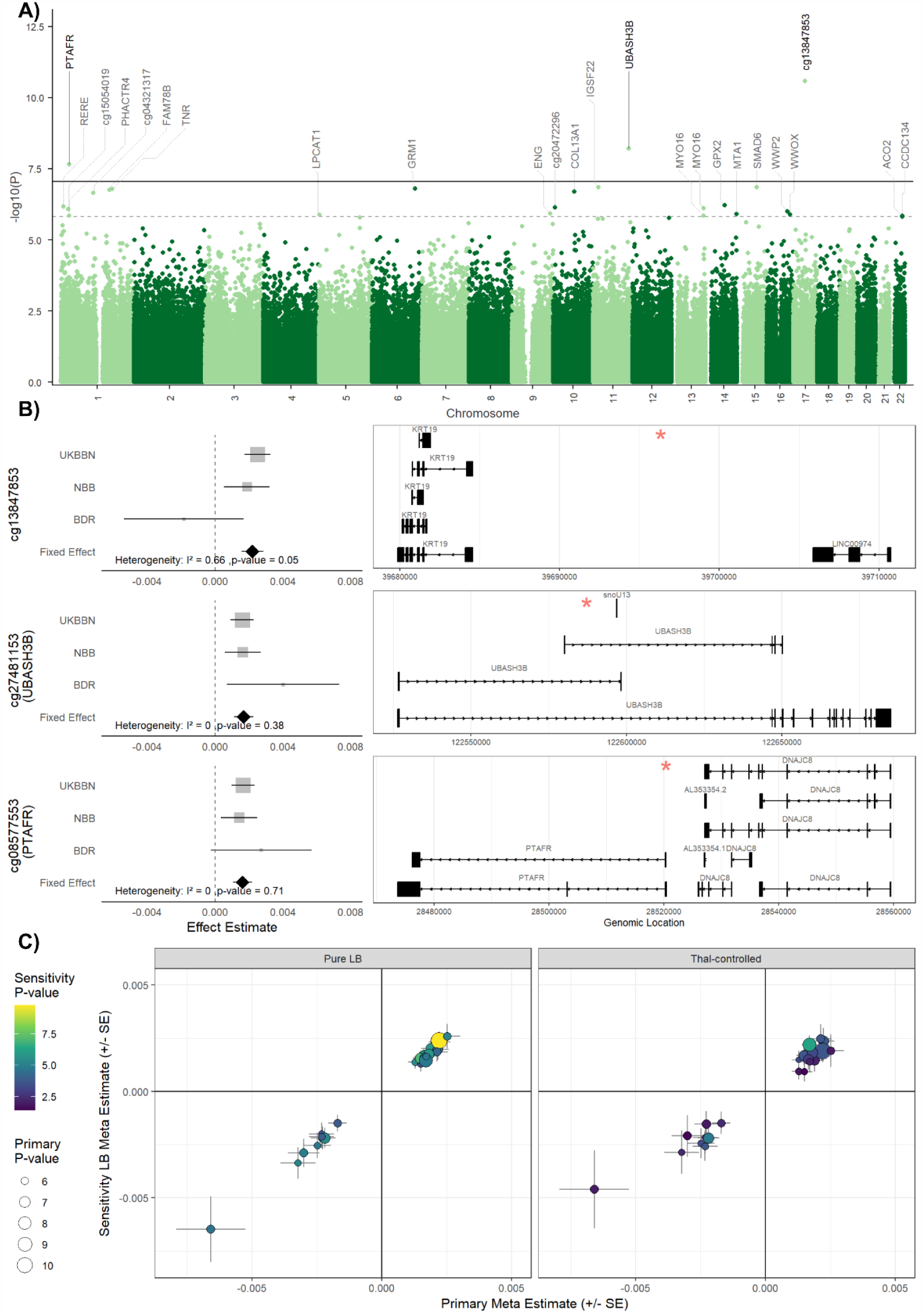
DNA methylation associated with Lewy body Braak stage in a cross cortical meta-analysis. **A)** Manhattan plot showing differentially methylated sites associated with Braak LB stage in a fixed effects inverse variance weighted meta-analysis of 1,217 total samples (primary full cohort LB pathology meta-analysis). Y-axis shows -log10 transformed p-value of association, x-axis shows chromosomal positions (chromosomes 1-22). The black solid horizontal line shows the genome-wide significance threshold (p < 9 x 10^-8^), whilst the grey dashed horizontal line denotes the FDR-corrected significance threshold (p_adj_ < 0.05). Points are colored to show chromosome number and for those that passed the significance thresholds these are annotated by UCSC gene symbols in the Illumina manifest file or, where unavailable, Illumina CpG ID is displayed. **B)** Forest plots of the effect sizes for the three sites passing the genome-wide significance threshold in the primary full cohort LB pathology meta-analysis. Grey points show cohort specific estimates, sized by weight in the inverse variance weighted meta-analysis. Whiskers show 95% confidence interval (CI) of estimate. Diamonds show effect estimates and 95% CI for fixed effect estimates from the primary meta-analyses. The I2 heterogeneity statistic and heterogeneity p-value are displayed for each site. Genome tracks for each methylation locus (Genome build GRCh37-hg19) are shown to the right, with the location of the associated methylation site marked with a red asterisk. **C)** Correlation plot of effect sizes between the primary full cohort LB pathology meta-analysis (y-axis) versus a subset analysis of samples without substantial coincident AD NFT pathology (“Pure LB”, Braak NFT stage ≤ II, left plot) or samples where Thal-staging was available to control for concomitant Aβ pathology (Thal-controlled, right plot) (x-axis) for the 24 FDR-significant DMPs identified in the primary full cohort meta-analysis. Points are sized by -log10 transformed p-value significance in the primary full meta-analysis and colored by -log10 transformed p-value significance in the pure LB subset analysis. Effect sizes are displayed as change in methylation (beta value) per increasing LB Braak stage.

Although we had controlled for Braak NFT stage in our primary meta-analysis, we wanted to further test if coinciding AD related co-pathology could be contributing to our findings. Two sensitivity analyses were performed, the first removing samples with a Braak NFT stage > II (“Pure-LB”, N = 763, 496 unique donors) and the second sub-setting the analysis to samples with Thal-staging present and including this as an additional covariate to control for Aβ pathology (“Thal-controlled”, n = 688 samples, 535 unique donors). In both instances, this reduced our total sample size and as a likely result, we observed a reduced number of DMPs passing either threshold for multiple testing correction. We observe one site in the Pure-LB analysis (cg13847853, p = 1.68 x 10^-10^, Effect = 0.0024, **Supplementary Table 3**) and one site in the Thal-controlled analysis (cg14888636, *MAN1C1*, p = 1.11 x 10^-8^, Effect = 0.0031, **Supplementary Table 4**) associated at the genome-wide significance threshold level. However, despite this, we found that the effect size of the 24 FDR significant DMPs we had identified in the primary full cohort analysis were highly correlated with the effect size from these secondary subset cohort analyses (Pure-LB: Pearson’s correlation (r) = 0.998, p = 1.64 x 10^-28^, Thal-controlled: Pearson’s correlation (r) = 0.982, p = 1.33 x 10^-17^), with a 100% concordance in the direction of effect in both cases (**Figure 2C**). Standard error estimates were also highly comparable between the primary and subset sensitivity analyses (Pure-LB: Pearson’s r = 0.998, p = 5.33 x 10^-29^; Thal-controlled: Pearson’s r = 0.99, p = 2.06 x 10^-20^). All sites prioritized in the primary meta-analysis also retained a nominal association (p < 0.05) in the secondary sensitivity analyses (**Supplementary Table 5**). Together this indicates that the associations identified in the primary full cohort analysis are not dependent on the presence of concomitant NFT or Aβ pathology, and that the loss of significance in this secondary subset analysis is principally a result of reduced power.

### Ontological enrichment analysis highlights neuronal and lysosomal processes and *SNCA* as a gene of interest

Next to determine potential biological function underpinning the methylation signature identified in our primary full cohort LB pathology meta-analysis, we employed pathway enrichment analysis, identifying a set of nine FDR-significant enriched terms (**Figure 3A, Supplementary Table 6A**). This included a number of terms relevant to neuronal and synaptic function (dendritic spine, regulation of nervous system process and regulation of postsynaptic membrane potential), inflammation (tertiary granule) and broader cellular processes (e.g., regulation of lipase activity, beta-catenin binding, regulation of protein-containing complex assembly, homotypic cell-cell adhesion and adherens junction). The KEGG pathway analysis revealed a single FDR-significant enriched pathway, annotated to lysosomes (**Figure 3B, Supplementary Table 6B**). Notably, despite not passing multiple testing correction, highly relevant terms to LB pathology were enriched at nominal significance, including endocytosis and PD.

**Figure 3.**
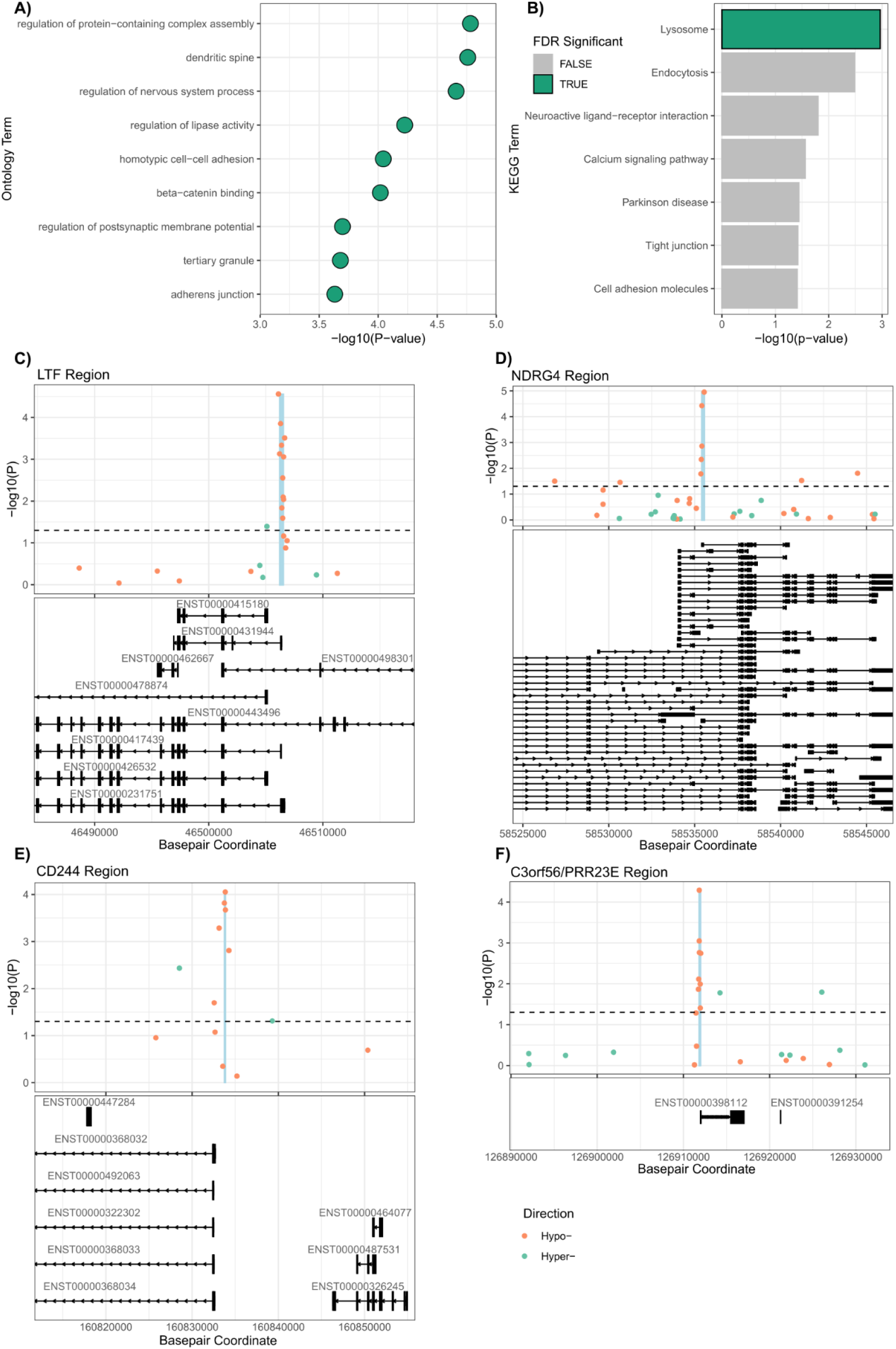
Pathway and regional analysis for the primary full cohort Lewy body pathology meta-analysis. **A)** Gene ontology (GO) analysis revealed nine FDR-significant terms, which are shown, with -log10 transformed raw p-value shown on the x-axis. REVIGO prioritized gene parent terms are shown on the y-axis. **B)** KEGG pathway analysis revealed one FDR significant term. Shown are all nominally significant enrichments, with the FDR significant term highlighted and with -log10 transformed raw p-values shown on the x-axis. **C-F)** Mini-Manhattan plots showing the four significant formal DMRs. Shown are **C)** the *LTF* gene region, **D)** the *NDRG4* gene region, **E)** the *CD244* gene region and **F)** the *C3orf56* gene region. The y-axis shows -log10 transformed p-value for the primary full cohort LB pathology meta-analysis and the x-axis shows the base-pair position. Points show probe coverage for the EPIC array and are colored by their direction of effect in association with LB Braak stage (hypomethylation = orange, hypermethylation = green). The highlighted blue region shows the differentially methylated region (DMR) identified from comb-p analysis. Below track shows the annotated transcripts for the region. The black dashed horizontal line represents nominal significance (p < 0.05).

The method employed for the ontological analyses, which aggregates methylation around gene annotations to provide a set of genes to be tested for over representation analysis using robust rank aggregation (RRA), identified 239 genes as significant based on the fixed-effect p-value from our primary full cohort LB pathology meta-analysis (**Supplementary Table 7**). Notably, this included the *SNCA* gene, which had not featured in the top results from our primary full cohort meta-analysis but is of considerable interest given it encodes for α-synuclein. Exploring this region in more detail, we saw that six loci in the *SNCA* gene showed evidence of a nominal association (uncorrected p < 0.05), all residing in the 5’ region of the gene and showing hypomethylation with increasing LB Braak stage (**Supplementary Figure 2**).

Next, we were interested in exploring whether *SNCA*, or indeed any other genes, contained statistically formally defined differentially methylated regions (DMRs), consisting of multiple adjacent DMPs by performing comb-p analysis on the summary statistics from our primary full cohort LB pathology meta-analysis. Although this analysis did not highlight the *SNCA* region as a formal DMR, we did identify four significant DMRs (**Supplementary Table 8**) overlapping the genes *LTF* (**Figure 3C**, Chr3:46506206-46506519, Šidák corrected p-value = 3.70 x 10^-7^), *NDRG4* (**Figure 3D**, Chr16:58535416-58535556, Šidák corrected p-value = 3.97 x 10^-6^), CD244 (**Figure 3E**, Chr1:160833741-160833863, Šidák corrected p-value = 1.27 x 10^-5^) and C3orf56 (**Figure 3F**, Chr3:126911830-126911953, Šidák corrected p-value = 9.77 x 10^-5^).

### Exploring cortical LB associated methylation changes in the context of other neurodegenerative diseases

Next, we sought to investigate the disease specificity of the LB associated DMPs we had identified in our primary full cohort LB pathology meta-analysis by comparing these results to summary statistics in previously reported EWASs in AD and Huntington’s disease (HD) brain samples. For AD associated effects, we utilized the summary statistics from a prior meta-analysis of NFT pathology in the cortex^12^, performed on the Illumina Infinium Methylation 450K array and encompassing data from 1,408 individuals. For HD associated effects, we used a recently generated case-control cohort of 41 individuals and encompassing matched EPIC array data from the entorhinal cortex (n = 38) and striatal (n = 37) brain regions^16^.

Taking the effect sizes for the Braak-LB associated DMPs from our primary full cohort LB pathology meta-analysis at a suggestive significance threshold (p < 1 x 10^-5^, n = 71 loci), we observed a significant overlap in the direction of effect with HD-associated changes in the entorhinal cortex (**Figure 4A&C, Supplementary Table 9A**, binomial test probability = 0.75, p_adj_ = 1.17 x 10^-4^), with 53 of the 71 CpGs assessed across both studies showing the same direction of effect. We found a nominally significant overlap with the direction of effect to Braak NFT associated changes for the 20 CpGs that overlapped between the studies (i.e., present across both array platforms) (**Figure 4A&B, Supplementary Table 9A**, binomial test probability = 0.75, p = 0.021, p_adj_ = 0.062), with 15 of the common 20 CpGs showing the same direction of effect. The lack of FDR significant overlap in this binomial test may in part be due to a lower number of overlapping CpGs between the EPIC and 450K array for testing. However, when we assessed the Pearson’s Correlation measure for direction of effect comparisons between the Braak-LB and Braak-NFT meta-analyses results, as prioritized by the nominally significant Braak-LB CpGs, we did observe a significant positive correlation (Correlation Coefficient = 0.51, p_adj_ = 0.002, **Figure 4B**).

**Figure 4.**
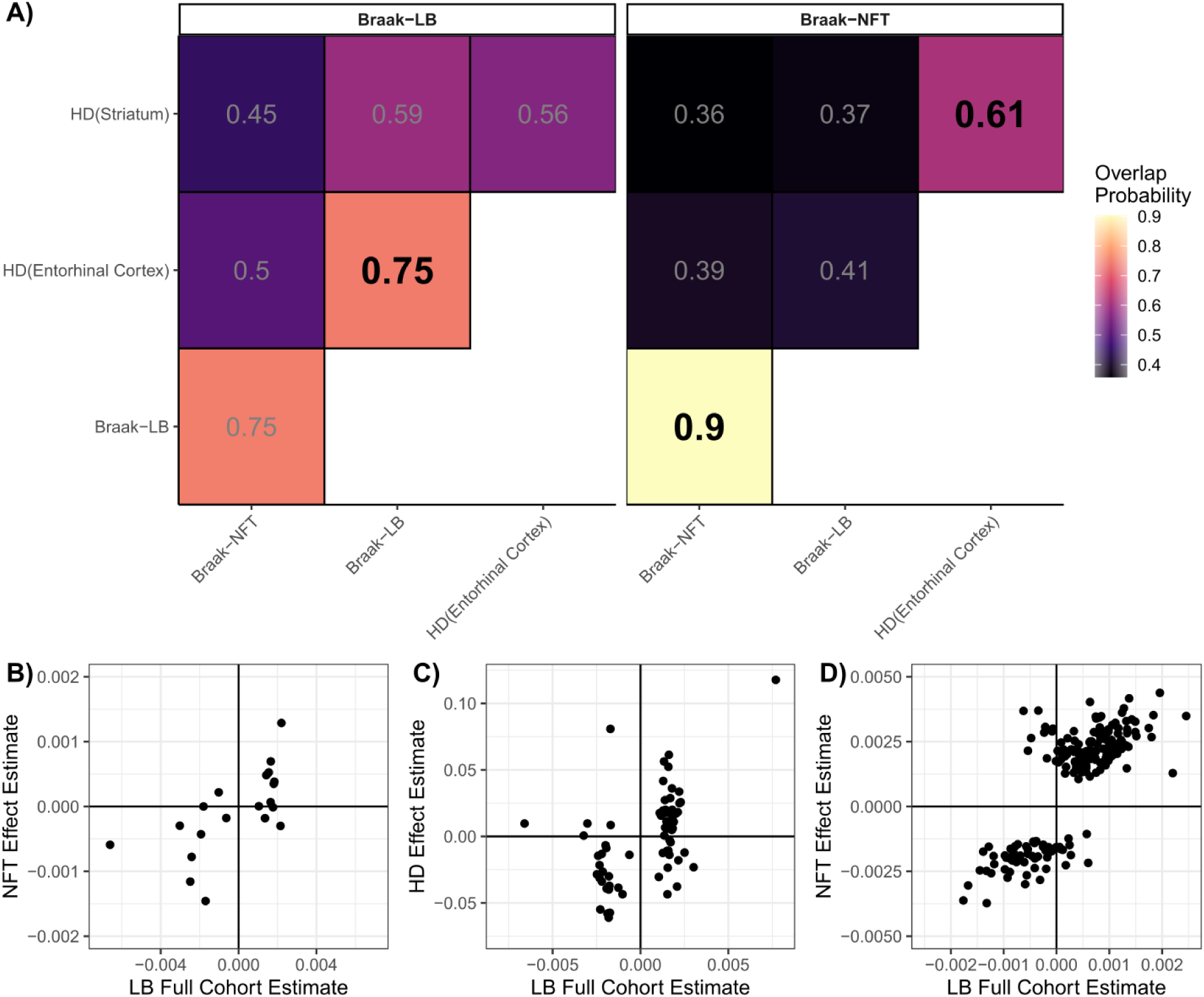
Summary of comparisons of epigenomic signatures between different pathologies. **A)** Heatmap of overlap in direction of effect for the most significant LB-Braak associated sites (p < 1 x 10^-5^, left panel) and NFT-Braak associated sites (p < 1.24 x 10^-7^, right panel) with the effect sizes for different neurodegenerative disease outcomes. The number displayed and fill color are the overlap probability for the same direction of association between two overlapping summary statistics as calculated by the binomial sign test. Bolded tiles indicate an FDR significant test result for significant overlap in directionality of associations. Braak NFT effects are taken from Smith et al. (2021) and HD effects from Wheildon et al. (2025). **B-D)** Scatterplots of DNA methylation associated with different pathologies. **B)** Shown for the top LB-associated loci is the comparison of effect size in cortex for LB and NFT (Smith et al., 2021) for the 20 overlapping sites across summary statistics, and **C)** for LB and HD (Wheildon et al., 2025) for the 71 overlapping sites across summary statistics. **D)** shows the comparison of effect size in cortex for previously reported NFT associated loci (Smith et al., 2021) in our LB samples for the 208 sites overlapping across summary statistics.

When we took the Bonferroni significant Braak NFT associated DMPs from the Smith et al study (p < 1.24 x 10^-7^, n = 208 loci present on the EPIC array), we found a significant overlap in the direction of effects to Braak-LB associated changes at these CpG sites (**Figure 4A&D, Supplementary Table 9B**, binomial test probability = 0.9, p_adj_ = 1.2 x 10^-34^), with 188 of these 208 sites showing the same direction of effect. However, we did not observe a significant overlap in the direction of effect for the 207 NFT-associated sites present in the HD study with HD-associated changes in the entorhinal cortex (**Supplementary Table 9B**). We did however observe a significant overlap in the direction of effect for the 207 CpGs in the HD entorhinal cortex and HD striatum samples (**Supplementary Table 9B**, binomial test probability = 0.61, p_adj_ = 0.00266), with 127 sites showing the same direction of effect. Notably, neither the Braak-LB DMPs, nor the Braak NFT DMPs showed any significant overlap in the direction of effect with the midbrain HD-associated effects in the striatum (**Supplementary Table 9A&B**), indicating effects are cortex specific. Together, this suggests a shared neurodegenerative methylation signal in the cortex across these three primary pathologies.

However, although a shared cortical methylation signature is apparent across neurodegenerative diseases from our comparison of direction of effects, it is worth noting that the top-ranked loci are distinct across the different diseases. Indeed, for the 24 FDR significant LB-associated DMPs in our primary full cohort meta-analysis, only cg13847853 was Bonferroni-significant in the Braak NFT meta-analysis performed by Smith et al. (2021). Outside of this overlapping locus, no other sites from the primary full cohort LB meta-analysis even showed suggestive significance (p < 1 x 10^-5^) in either the Braak NFT or HD EWASs (**Supplementary Table 10**) and similarly, aside from cg13847853, none of the Bonferroni DMPs from the Braak-NFT meta-analysis showed suggestive significance (p < 1 x 10^-5^) in the primary full cohort LB pathology meta-analysis, nor in the HD EWASs (**Supplementary Table 11**). This indicates that the most robust cortical DMPs in each neurodegenerative disease are specific to that disease.

### Methylation quantitative trait loci analysis prioritizes sites under genetic influence but does not support overlap with known genetic risk loci

A number of studies have shown that DNA methylation can be influenced by genetic variation, commonly termed methylation quantitative trait loci (mQTLs). Given the size of our cohort, we utilized matched genotyping array data for 405 samples from the UKBBN cohort to test for mQTLs at the 24 FDR significant methylation sites nominated in our primary full cohort LB pathology meta-analysis. We identified 534 cis-mQTLs annotated to nine methylation loci: cg09966204 (*MYO16*), cg15054019, cg07837822 (*MYO16*), cg27625119 (*LPCAT1*), cg22516725 (*WWOX*), cg16262110 (*MTA1*), cg21912448 (*RERE)*, cg05523624 (*GRM1*) and cg25194322 (*FAM78B*) (**Supplementary Table 12, Supplementary Figure 3**). We did not identify any significant trans-QTLs. However, none of the SNPs corresponding to the identified cis-mQTLs overlapped with GWAS hits reported in summary statistics from GWAS of AD^17,18^, PD^4^ or LBD ^5^.

### The *SNCA* region shows enrichment of differentially methylated loci and evidence of colocalization with genetic risk signatures

To test if genomic regions prioritized by GWAS of AD^18^, PD^4^ and LBD^5^ show enrichment for LB pathology associated methylation, we employed Brown’s method to combine p-values (from the primary full cohort LB pathology meta-analysis) across the disease-associated linkage disequilibrium (LD) regions (**Supplementary Table 13**). Across the combined 99 regions from the three GWASs tested, only the *SNCA* region, as annotated in the recent LBD (Chr4: 90743331 – 91005096, p_adj_ = 0.0099) and PD (Chr4: 89835093 – 91453046, p_adj_ = 0.015) GWASs showed significance after multiple testing correction. This included the seven CpG sites that we prioritized after the RRA analysis.

Given our findings of enriched epigenetic alteration in the *SNCA* region, we next tested if the methylation sites in the region show evidence of QTL relationships with genetic variation in this region using the UKBBN cohort. We identified 16 methylation sites in this region that showed significant evidence of a cis-mQTL relationship including six of the CpG sites nominally associated with LB pathology in the primary methylation meta-analysis (**Supplementary Table 14**). Notably, the strongest mQTL relationship observed was to cg01966878 (**Figure 5A-B**), which was the most strongly associated site overlapping *SNCA* in the primary full cohort LB pathology meta-analysis (p = 1.67 x 10^-4^, Effect Estimate = -0.0034).

**Figure 5.**
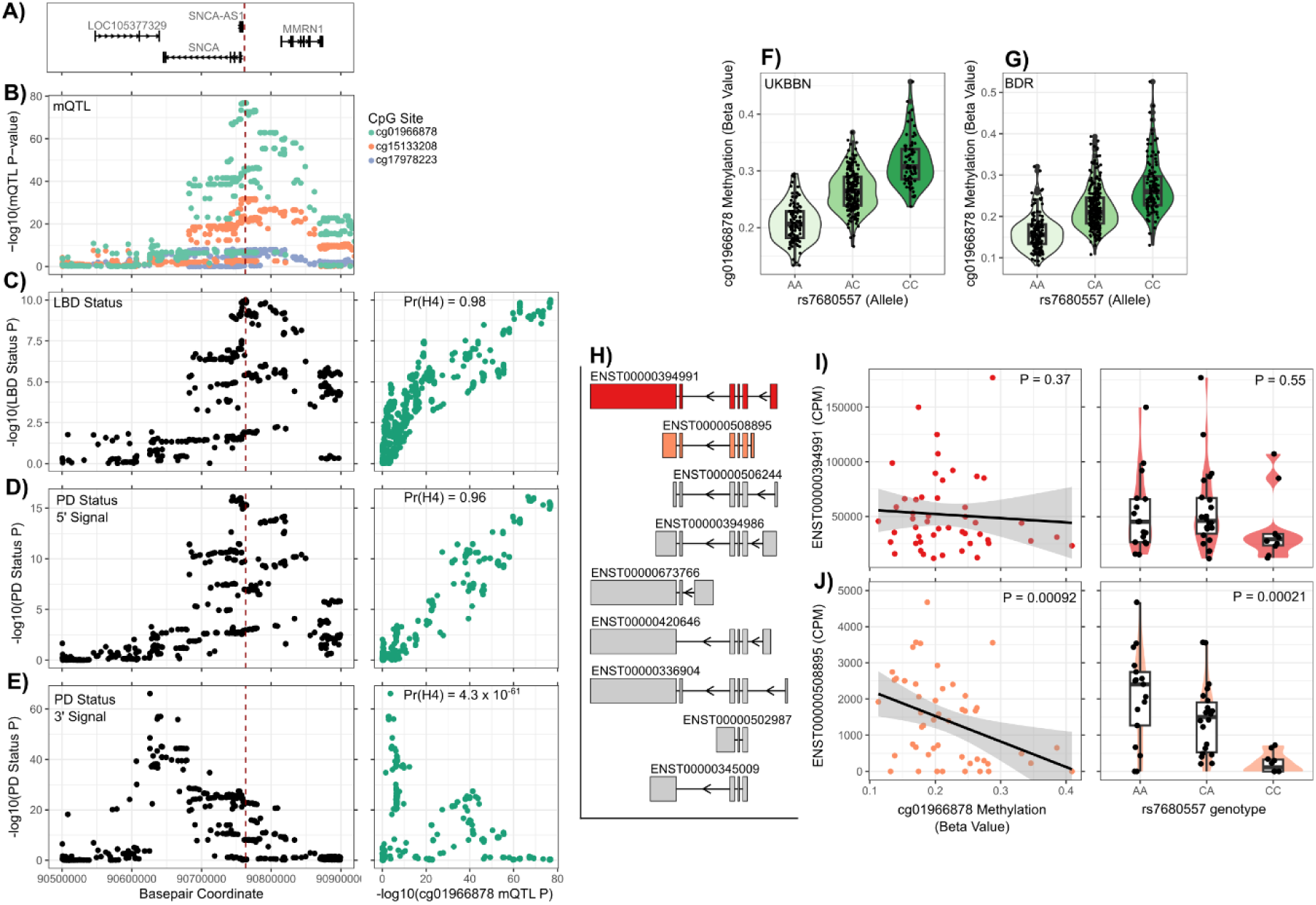
Characterization of *SNCA* methylation quantitative trait loci. **A)** Genomic tracks showing the *SNCA* coding region and surrounding genes (Build 37, hg19). Shown below the track are the same genomic regions with mini-manhattan plots showing the associations of the following traits: **B)** mQTL associations for the three methylation sites with evidence of colocalization with LB-relevant GWAS. Points are colored by cpg annotation for mQTL association, **C)** GWAS for Lewy Body Dementia (Chia et al. 2021), **D)** Conditioned GWAS for Parkinson’s Disease (PD) (Pihlstrom 2018), results represent associations controlling for the lead SNP in the 3’ *SNCA* region (rs356182). **E)** Conditioned GWAS for PD (Pihlstrom et al. 2018), results represent association controlling for the secondary associated signal in 5’ *SNCA* region (rs763443). On the right of each GWAS plot is a correlation plot displaying the -log10 transformed p-values for that GWAS on the Y-axis and the -log10 transformed p-values for the strongest mQTL signature (annotated to cg01966878) on the X-axis. The colocalization prior probability (Pr(H4)) is shown within each plot, with a value closer to 1 indicative of high probability of colocalization under a shared causal variant. Within each mini-manhattan plot is a red-dashed vertical line annotating the lead-SNP location for LBD (rs7680557). **F-G)** Box and violin plots showing lead-LBD SNP rs7680557 genotype and DNA methylation at cg01966878 in the UKBBN and BDR datasets. **H)** Tracks representing all called transcript isoforms of *SNCA* identified in the previously generated dataset in BDR (Leung et al, 2024). Highlighted in red is the highest expressed canonical transcript of *SNCA* (ENST00000394991) and highlighted in orange is the genotype and methylation associated alternative isoform (ENST00000508895). Shown to the right is the association with lead-LBD SNP rs7680557 and methylation at cg01966878 for **I)** the canonical and **J)** alternative transcripts. P-values represent linear association model results.

We next tested if the mQTL profiles in the *SNCA* region and the genetic association signatures of GWAS for LBD and PD show evidence of colocalization under shared causal variants. The mQTL signal for cg01966878 showed strong evidence of colocalization to both LBD genetic risk (**Figure 5C**, Colocalization Pr(H4) = 0.98) and the 5’ secondary conditional PD genetic signature (**Figure 5D**, Colocalization Pr(H4) = 0.96), but not to the strongest PD genetic risk signature located 3’ of the gene (**Figure 5E**, Colocalization Pr(H4) = 4.3 x 10^-61^). Evidence of colocalization was also observed at two additional CpG sites (cg15133208 and cg17978223, **Supplementary Table 15**). We observed that the most significant mQTL relationship of cg01966878 to the LBD risk locus rs7680557 identified in UKBBN (**Figure 5F**, Effect Estimate = 0.053, p = 1.63 x 10^-77^) is also reproduced in the BDR cohort (**Figure 5G**, Effect Estimate = 0.054, p = 7.39 x 10^-69^). We highlight that the direction of effect between the SNP and DNA methylation at cg01966878 is concordant with the expected direction from the original GWAS and methylation meta-analysis findings. That is, the A-allele, which is associated with increased risk of LBD, is strongly associated with hypomethylation, which in turn is the direction we observed in association with increased Braak LB-stage (**Supplementary Figure 2**).

Using available reference data from the ROSMAP cohort^19^, we tested the association of both the genetic locus and the top prioritized methylation locus on *SNCA* gene expression. When assessing short-read RNA sequencing data, we find no evidence of an effect of either rs7680557 or cg01966878 on *SNCA* transcript expression (**Supplementary Figure 4**). Despite a previously reported expression QTL relationship with the *SNCA*-antisense (*SNCA-AS*1) transcript, we do not observe evidence of association of rs7680557 or cg01966878 to *SNCA-AS1* expression (**Supplementary Figure 4**), although the expression of *SNCA-AS1* in this dataset was notably very low. Although, we had not observed an association at the overall transcript level, we next tested if splicing isoform associations could be detected with rs7680557 or cg01966878 by leveraging a long-read RNA sequencing dataset of 49 samples that had been previously generated in the BDR cohort^20^ (**Figure 5H**). A *SNCA* transcript resulting in differing 5’ and 3’ untranslated region (UTR) structure (ENST00000508895) was significantly associated with both the LBD risk variant rs7680557 (**Figure 5J**, p = 0.00021) and the mQTL prioritized cg01966878 methylation site (**Figure 5J**, p = 0.00091). Consistent with the short-read RNA sequencing data, we did not observe any evidence of association with the canonical primary transcript isoform for *SNCA* (ENST00000394991) and either rs7680557 (**Figure 5I**, p = 0.55) or cg01966878 (**Figure 5I**, p = 0.37).

## DISCUSSION

In this study we present a meta-analysis of cortical DNA methylation associated with LB pathology. We find evidence of significant differential methylation at 24 loci across the genome, the top three of which reached the most stringent cutoff for genome-wide interpretation (i.e., cg27481153 (*UBASH3B*), cg08577553 (*PTAFR*) and cg13847853. We find evidence that the functional annotations enriched in prioritized loci are related to a number of brain relevant traits including neuronal and synaptic function. In keeping with known pathway relevance in LB-disease, we also find evidence of enrichment for lysosomal processes in our results. Comparison of our results to previous EWASs of AD-related neuropathology (Braak NFT) and HD status indicates a shared epigenomic profile across some, but not all, neurodegenerative diseases. Notably we observe comparable cortical profiles of LB-pathology, HD-status and NFT-pathology, but minimal comparability between AD and HD profiles. When assessing known GWAS regions associated with LB-diseases, we find evidence of hypomethylation in the *SNCA* region and when looking at mQTL relationships, find evidence of strong colocalization with disease associated genetic variation located at the 5’ region of the gene. We further provide evidence that these changes associate with the expression of an alternative *SNCA* transcript isoform, but not the primary canonical transcript.

A number of the genes prioritized from the primary full cohort LB meta-analysis have been previously related to LB-related traits. *UBASH3B* encodes Ubiquitin Associated and SH3 Domain Containing B, which is highly expressed in brain tissue and has a variety of related functions in cell signalling^21^. It has been previously associated in PD and DLB contexts, reported as differentially expressed in single cell analyses of prefrontal cortex astrocytes^22^ and inhibitory neurons of the anterior cingulate cortex^23^. Rare variants in the gene have also been associated with serum total tau levels in an exome sequencing study^24^. *PTAFR* encodes Platelet Activating Factor Receptor, highly expressed in brain tissue and implicated in cell signaling via the NF-κB pathway. It has been reported to be differentially expressed in AD hippocampal tissue^25^ and is implicated in mediating astrocytic inflammatory responses^26^. cg13847853 is a more perplexing site in terms of functional annotation but has been previously associated with AD NFT related pathology^12^. It resides between a long non-coding RNA (*LINC00974*) and the gene *KRT19*. *KRT19* is minimally expressed in brain, although it has been associated on a proteomic level in the periphery to Aβ pathology^27^. Further work is required to elucidate the downstream regulatory impact of this methylation site.

Of the FDR significant differentially methylated loci, a number have been previously associated with parkinsonian and LB related traits. *WWOX*, encoding WW Domain Containing Oxidoreductase, is a gene which is linked to autosomal recessive neurodevelopmental disorders^28^, but has also been shown in large GWAS to be associated with both AD status^18^ and the development of cognitive decline in PD^29^. We note that an FDR-significant site annotated to the gene *GRM1*, encoding glutamate metabotropic receptor 1 also lies downstream of the gene *RAB32* which has been recently identified as a causal gene in autosomal dominant PD^30^.

To address the potential for confounding AD related NFT pathology, we utilized a secondary meta-analysis restricted solely to samples with minimal co-pathology as defined by Braak NFT staging. This reduced the total viable sample size and, perhaps unsurprisingly, resulted in far fewer sites passing multiple testing correction. Solely cg13847853, the topmost associated loci in our primary full cohort LB pathology meta-analysis, retained genome-wide significance. Similarly, when we restricted the analysis to samples with Thal staging information available and controlled for this as a covariate in meta-analysis as a method of addressing Aβ related pathology, only a novel locus at *MAN1C1*, not detected in our primary full cohort LB pathology meta-analysis, reached genome-wide significance. Notably however, there was a striking concordance in estimated effects between the primary and sensitivity meta-analyses, which we interpret as evidence that the LB-stage associated effects from our primary analysis are not contingent on the presence of coincidental NFT and Aβ pathology.

A number of the post-hoc tests on our results implicate epigenetic changes in the *SNCA* gene, despite this region not passing the multiple testing threshold used in our primary or secondary meta-analyses. In a targeted assessment of the gene, we found evidence of hypomethylation in a number of sites in the 5’-region of the gene, which we believe warrants highlighting given that multiple other studies have reported the same phenomena in relation to LB disease^31–35^. Furthermore, when employing a method to aggregate p-values from our meta-analysis results based on genetic regions reported from related GWAS, we found that the only significantly enriched region corresponded to the *SNCA* region reported in the most recent PD^4^ and LBD^5^ GWASs. We find evidence that a number of methylation loci at the 5’ region of the gene shows strong evidence of mQTL relationships. The most significant mQTL relationship observed in this region showed evidence of colocalization and shared causal variants to the genetic risk profile of LBD and the secondary, conditioned genetic risk signature of PD. This observation is in line with previous work^36^ and provides evidence that genetic variation in the 5’ region of *SNCA* is relevant to functional alterations in the cortex, in-keeping with findings in relation to both LBD and REM sleep behavior disorder^5,37^. Although we do not find evidence of expression alterations in *SNCA* or *SNCA-AS1* resulting from implicated genetic or epigenetic loci, we do report evidence of isoform specific associations in *SNCA*. Specifically, ENST00000508895 was associated with both the lead genetic risk locus for LBD and our identified mQTL. This transcript does not result in alterations in the peptide sequence of *SNCA* but is altered in its 5’ and 3’ UTR compared to the canonical *SNCA* transcript isoform. ENST00000508895 has also been previously reported as an LBD-risk specific QTL in an independent dataset^38^. More work is warranted to determine the functional implications of this transcript. Given that *SNCA* differential methylation has a reported cell-type specific effect^39^, future studies looking at cell type specificity of DNA methylation in *SNCA* may yield sites in the gene at genome-wide significance.

We further find evidence of a shared epigenomic signature across multiple neurodegenerative diseases and pathologies. There was a clear comparability between epigenetic effects for LB and NFT pathologies in the cortex, indicating a level of shared molecular processes related to both pathologies. However, notably only cg13847853 was associated at genome-wide significance in both analyses, indicating that despite a shared profile between the two, the prioritization of individual processes may differentiate the different pathologies. Indeed, this indication of a level of unique signature to either pathology is displayed in our exploration of epigenomic prediction models, where best balanced accuracy was achieved only when using a model trained for the specific pathology being predicted. This shared effect is consistent with other modalities of genetic and biological data, with genetic risk in genes such as *CLU*, *WWOX* and *GRN*^40^, as well as the *HLA* region^41^ having been shown to have associated overlap between PD and AD and evidence of a level of shared transcriptomic alteration in DLB and AD brains^42^. Perhaps more surprisingly was evidence found here between LB and HD associated epigenomic profiles, specific to the cortex. This shared effect did not appear to be indicative of a broad neurodegenerative signal in the cortex, as no evidence of comparability was seen between HD and NFT pathological profiles. Similarly, we did not find evidence of shared profiles with HD associated changes in the striatum. Given that HD is caused by a highly penetrant autosomal dominant genetic defect and shares minimal pathology with LB disease, this finding is perplexing. One potential explanation is that both diseases are marked by a shared degeneration of the striatal regions and the similar cortical epigenomic profiles may represent a secondary signature of midbrain neurodegeneration. Another explanation may be that as the disease progresses, HD cortical profiles show similar underlying epigenetic profiles to those observed in the vulnerable striatal regions. Further work is needed to determine if this shared epigenomic profile we observed between these differing movement disorders represents a potential shared biological etiology or secondary effects to other common features of the two conditions.

We found evidence that some of the FDR significant methylation sites identified in our primary meta-analysis have evidence of genetic influence from our mQTL calling. However, we found no evidence that the genetic variants prioritized from this analysis are associated with neurodegeneration related phenotypes from previous GWAS. This is in-keeping with similar findings from previous meta-analyses of neurodegenerative pathologies which have failed to find evidence of pleiotropy between AD genetic risk variants and mQTLs associated with primary EWAS prioritized methylation sites^12^.

One limitation of our study is the quantified neuropathological outcomes used to determine association and control for confounding covariance. In this analysis we have used Braak LB stage, due to its use in previous studies of this type both for LB disease and AD^13^. Although Braak LB stage is a useful instrument in assessing PD-relevant pathology progression, its utility in the broad context of LB diseases, in particular DLB, is highly questioned. Indeed, alternate staging schemes focus more on categorically subtyping pathological LB by its distribution in different brain regions^43^ as opposed to an ordinal semi-quantitative scheme that assumes pathology initiates in the brainstem. Thus, in our analysis, samples which are grouped into specific Braak LB stages may present highly variable LB pathology burden in the cortical regions we are assessing. This decision was chosen for practicality to allow for a harmonized measure between this study and previous EWASs. Braak LB staging was also the most represented pathological staging scheme in the brain bank records that we had access to. Similarly, although we made efforts to stratify and control for AD co-pathology as measured by Braak NFT stage as this was available for all samples, we could not control for Aβ pathology for all samples used. This is because measurements of Aβ deposition, such as the Thal stage, were absent for a large proportion of available cases. We did seek to address this potential confounder when we performed the lower powered sensitivity EWAS that included Thal stage as a covariate however, further work is needed to fully determine the effect of Aβ on LB-associated epigenomic profiles. One challenge when studying post-mortem brain tissue in neurodegenerative diseases is the impact of neuronal cell loss, and gliosis on cell populations, which may impact the DNA methylation signatures observed in disease. Although we have attempted to mitigate this by controlling for the proportions of different neuronal and glial cell types, future analyses should be undertaken on sorted cell populations in order to identify cell-type specific signatures. Differences in the cell type deconvolution model used in this study, which resolves five cellular proportions, compared to less-granular models which resolved fewer cell types employed in previous studies, may also account for differences in results with previous EWASs in LBDs^13^. Notably, a recent study from our group testing our LB meta-analysis results with a novel tool for measuring enrichment of cell specific epigenetic signatures reported enrichment of our LB DNA methylation signature in microglial and oligodendrocytic cells^44^.

In summary, we have undertaken the largest meta-analysis of DNA methylation in LB diseases to date, highlighting robust methylation alterations at 24 loci, and highlighting epigenetic alterations in *SNCA* in disease. Future studies warrant DNA methylomic analyses of well-characterized cohorts with detailed ante-mortem and post-mortem clinical and pathological assessments, in order to identify signatures that distinguish between different LBDs that show similar pathological profiles with more distinct clinical symptoms. The integration of DNA methylation data with other epigenetic marks and transcriptional profiling will allow the identification of disease signatures that impact at multiple layers of genomic regulation and may represent novel candidates when developing pharmacological intervention strategies.

## METHODS

### Cohort descriptions

Samples were sourced from the UK Brain Bank Network (UKBBN), an online database covering brain bank centers across the UK and annotated with demographics, neuropathological and clinical assessments. All available records on the UKBBN were downloaded (June 2019) for manual selection. First, to determine LB positive case availability, records were filtered on the basis of neuropathological diagnostic criteria. Only cases with an annotation of Braak LB stage of 3-6, or annotation of neocortical or limbic LB pathology were selected as LB disease cases. The majority of samples matching our search criteria were found in brain banks at Oxford, Newcastle, Imperial College London and University College London. Samples were sourced on the basis of a primary diagnosis of either PD or DLB, either in available clinical records or at post-mortem examination. Samples were included if they had a primary neuropathologically confirmed diagnosis of PD or DLB and, where primary diagnosis was absent, had a “High” assessment of the likelihood that the pathological profiles were associated with typical DLB clinical syndrome from the 2017 Lancet criteria^2^. Exclusions were made for familial synucleinopathies or misdiagnosed primary symptoms (e.g., multiple system atrophy (MSA), Progressive Supranuclear Palsy (PSP) etc.), severe cerebrovascular disease or TDP-43 pathology, as well as removing cases annotated with amygdala predominant LB pathology. Seventy-one cases had annotation for LB disease relevant pathology following the 2009 Unified staging scheme but lacked Braak LB staging annotation^45^. To harmonize these annotations for analysis alongside the Braak scheme, records were converted in accordance with the BrainNet Europe Consortium^46^. Sixty-one cases with “Neocortical Lewy Body Disease” were annotated as Braak LB stage 6. The remaining ten cases showed evidence from the available additional neuropathology notes of LB pathology in the temporal cortex or cingulate gyrus and were annotated as Braak LB stage 5. From this criterion we selected 28 cases with Braak LB stage 3, 38 cases with Braak LB stage 4, 39 cases with Braak LB stage 5 and 211 cases with Braak LB stage 6. An additional 90 control individuals without neurological disease were sourced, which were chosen to exclude significant cerebrovascular pathology or substantial AD-related pathology (Braak NFT stage > II). A set of 20 samples lacked explicit Braak NFT stage in their record but had sufficient tau staining reported in the neuropathology records for Braak NFT stage to be imputed. In total, a set of 417 total donors from UKBBN were chosen for our study, with two brain regions sourced for the majority of individuals. These brain regions were the ACC (Brodmann Area (BA) 24) and the PFC (BA9). All samples received for our study had informed consent at local brain banks, covered under individual ethical agreements at the center (REC references 24/NE/0012, 15/SC/0639, 18/LO/0721, 23/WA/0273). Ethical approval for our study was granted from the University of Exeter Medical School Research Ethics Committee (13/02/009).

Data for the Netherlands Brain Bank (NBB) cohort has been previously detailed in Pihlstrøm et al., 2022, where they had analyzed PFC tissue from 73 control, 29 incidental LB disease, 139 PD (of which 74 were PDD), and 81 DLB patients^13^. Data from the Brains for Dementia Research Cohort (BDR) was previously detailed in Shireby et al., 2022^14^. Neurological controls were selected with absence of LB-pathology and low AD co-pathology (Braak NFT < III), along with the absence of significant neuropathological co-pathology (TDP-43 pathology, gross infarction, severe cerebrovascular disease). Eight samples overlapped between the UKBBN and BDR cohort and were removed from the BDR cohort prior to analysis. Case samples were selected in keeping with the same criteria as the UKBBN. A final set of 116 samples from the BDR cohort were used for the meta-analysis.

### Sample preparation and methylomic profiling

For the UKBBN cohort, tissue pieces ranging between 25-35 mg were cut from frozen PFC or ACC tissue before being milled over liquid nitrogen. Following tissue grinding DNA was extracted using the Qiagen AllPrep DNA/RNA/miRNA Universal kit according to the manufacturer’s instructions and stored at 4°C. 500ng of DNA from each sample was bisulfite treated using the Zymo EZ-96 DNA methylation Gold kit. Methylomic profiling was completed after sample randomization using the Illumina Infinium MethylationEPIC v1.0 BeadChip.

### DNA methylation analysis, quality control and normalization

Data processing was performed using R, and signal intensity files were loaded into R using the minfi package^47^. All data was subjected to a thorough quality control (QC) pipeline using the minfi^47^ and wateRmelon packages^48^. A MethylumiSet object was created from iDATs using the readEPIC function from wateRmelon^48^ and an RGChannelSet was created using the minfi package^47^. Samples were excluded from further analysis for the following reasons: (1) they had a median methylated or unmethylated sample intensity < 1,500 or < 1,000, respectively, (2) that the bisulfite conversion efficiency was < 80%, (3) there was a mis-match between reported and predicted sex, (4) if the sample was determined to be an outlier using the outlyx function or, (5) if the samples were found to be cryptically related to one another or unrelated to its matched sample from the other brain region. Finally, using the pfilter function in the WateRmelon package^48^ samples with a detection p-value > 0.05 in > 5% of probes, probes with < three beadcount in 5% of samples and probes having 1% of samples with a detection p-value > 0.05 were also removed. Following QC, samples not passing the aforementioned checks were removed and quantile normalization was performed using the dasen function in the wateRmelon package^48^. Beta values were extracted for further analysis. Cell type proportion calculations were implemented using the projectCellTypeWithError() function within the CETYGO package^49^, using the reference panel resolving enriched profiles for GABAergic neuron, glutamatergic neuron, oligodendrocyte, microglia and astrocyte reference data^50^.

### Within cohort epigenome wide association assessment

Prior to meta-analysis, the association with LB Braak stage was assessed within each cohort in a harmonized approach. Within each cohort, linear models were fit using the base lm() function in R to test for associations between LB Braak stage and DNA methylation at each site of the genome. This is displayed by the following equation for the NBB and BDR analyses:

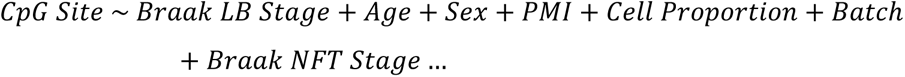

For the UKBBN analysis, given two brain regions were present per individual, this caused a violation of linear regression by non-independence between samples type. To correct for this, a mixed effect model was employed using the lme() function within the nlme package. Brain region was included as a fixed effect covariate and individual ID was supplied as a random-intercept covariate to account for paired observations. This can be displayed in the following formula:

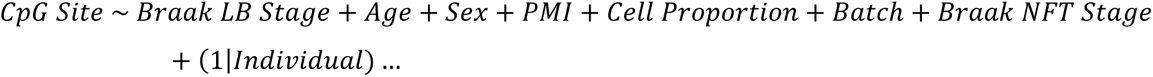

Consistent covariates of age, sex, post-mortem interval (PMI), cell proportions and Braak NFT stage were included in all three cohorts. For the UKBBN and BDR cohorts, brain bank source and array plate batch were included as processing batch variables. For a small subset of the UKBBN donors, PMI values were missing (n = 12) and in these cases, mean imputation was used to fill missing values. This method was justified based on previous data exploration for best methods to deal with missingness in phenotypic data^51^. For NBB, only processing plate batch was included as an additional covariate. For each cohort, model p-value inflation was assessed genome-wide and surrogate variables added to reduce the lambda value below a 1.2 threshold. One surrogate variable was included for BDR and three were included for NBB, with no surrogate variables required to reduce inflation in UKBBN (**Supplementary Figure 1**).

### Meta-analysis

Estimated regression coefficients and standard errors were extracted from each model and used for meta-analysis of all probes overlapping each dataset (n = 774,310). Harmonized models were meta-analyzed using the metagen function in the meta R package^52^, as had been employed for previous EWAS meta-analyses^12,13,53^, which applies an inverse variance weighting. Both fixed and random effects were assessed, with the fixed effects model being the primary interpreted results, given the number of cohorts available and the observation that the majority of top sites identified did not show evidence of significant heterogeneity (**Supplementary Tables 2-4**). In addition to fixed and random effects modelling statistics (Effect estimates, standard error and p-values), the I2 heterogeneity measure and corresponding p-values were also extracted.

For interpretation of results, a strict genome-wide significance threshold was based on a previous simulation and permutation testing study to define a significance level as determined by Mansell and colleagues^54^. This approach sets a consistent significance threshold of p-value < 9 × 10^-8^ for any study utilizing the Illumina EPIC array and was determined based on multiple permutations of null EWAS study scenarios to determine a 5% family wise error rate. A less stringent multiple testing False Discovery Rate (FDR) significance threshold (p_adj_ < 0.05) was also used in our study. Finally for more exploratory post-hoc analysis, a more lenient suggestive threshold was applied of uncorrected p-value < 1 x 10^-5^.

### Differentially methylated region analysis

To identify differentially methylated regions (DMRs) from summary statistics from our meta-analyses, we employed the combined p-values method as implemented in the python package comb-p^55^, which identifies DMRs based on a defined sliding window, correcting for spatial autocorrelation between adjacent methylation sites. We performed this with the default settings for consistency with prior meta-analyses, with a distance of 500 base-pairs (bp) and a seeded p-value of 1 x 10^-4^. Significant DMRs were defined as those with a Šidák’s multiple testing correction p-value < 0.05 and with ≥ two methylation sites residing in the identified DMR. For plotting, EnsDb.Hsapiens.v75 and GenomicRanges were used to extract annotated transcripts for genome build 37 hg19.

### Gene ontology analysis

Gene ontology (GO) analysis was performed using the package methylGSA function methylRRA^56^. This method assesses genome-wide summary statistics as summarized by p-values and aggregates them around gene annotations, controlling for the number of CpGs annotated to differing genes. We utilized the overrepresentation analysis approach, which applies a within gene Bonferroni correction and an across gene FDR correction to prioritize genes for ontological enrichment analyses. Given the overlap of similar ontology terms, for interpretation of significant enriched terms in our results, we further employed the REVIGO method, as implemented in the rrvgo R package^57^ using default parameters to identify shared parent terms for closely related GO annotations.

### Cross disease epigenomic signature comparison

Assessment of overlap between EWASs was conducted by assessing the directionality of estimated effects in available summary statistics. To statistically compare shared direction of effects, a one-sided binomial sign test was performed to test for greater than expected overlap in association directionality. For comparison to AD related Braak NFT associated signatures, summary statistics were sourced from Smith et al. 2021, which had performed a cross-cortical meta-analysis^12^. This had utilized 1,408 unique donors, which had been profiled on the Illumina Infinium Methylation 450K BeadChip. For assessment of HD-associated signatures, summary statistics were sourced from Wheildon et al. (2025)^16^. This included matched data for 38 entorhinal cortex and 37 striatal samples, profiled on the Illumina EPIC array. The Braak NFT EWAS signature comprised the 208 sites on the EPIC array corresponding to the 220 sites passing Bonferroni significance in Smith et al. (2021). For the Braak LB signature, given that more modest p-values were identified, we used the 71 sites passing our suggestive p-value threshold of 1 x 10^-5^ in the primary full cohort LB pathology meta-analysis. Following all comparisons tested, FDR correction was applied for interpreting the significant overlap in shared direction of association.

### Methylation quantitative trait loci analysis

Methylation quantitative loci (mQTLs) were identified in the UKBBN data using a set of 473 samples with matched Illumina Global Screening Array (GSA) genotyping data. mQTLs were tested for the 24 loci at FDR significance in the primary full cohort LB pathology meta-analysis. The R package matrix eQTL was used to identify methylation sites with significant association to genetic loci^58^. Methylation beta-values were tested against genotype (coded additively) utilizing a linear model for association testing while adjusting for confounders of age, sex, cell-type type proportion estimates and the first three genetic principal components. For the *SNCA* regional analysis, 36 methylation sites annotated to *SNCA* were subset and run as a separate analysis using identical methods to those described above for the 24 FDR nominated loci. Matrix eQTL was run in linear model mode with an additive genetic model to assess cis- and trans-mQTLs. Cis-mQTLs were defined as SNPs located within ±1 Mb of the associated CpG site, while trans-mQTLs correspond to SNPs > 1 Mb from the CpG. The significance threshold was adjusted for multiple testing using FDR correction, with a significance threshold of p_adj_ < 0.05.

### Genomic enrichment analysis

To test for genomic enrichment in LB associated methylation loci in known regions prioritized by prior GWAS, we employed Brown’s method, as implemented in the EmpiricalBrownsMethod R package to aggregate p-values across multiple methylation loci^59^. Regions in linkage disequilibrium (LD) with primary loci in GWAS were extracted for AD, PD and LBD. For PD ^4^ and AD^18^, reported regions were taken directly from the supplementary material of the respective study. For LBD^5^, as regions were not directly reported, regions were manually annotated based on the minimum and maximum position of variants at genome-wide significance (p < 5 x 10^-8^) in the genome-wide summary statistics around the five primary loci reported in the study. P-values were FDR corrected to account for the number of total genomic regions tested. Methylation beta matrices were z-score normalized using the scale() function in R within each cohort, to allow for the correlation calculations within the Brown’s methodology accounting for inter-cohort variability whilst retaining the inter-site correlations.

### Colocalization analysis

We utilized the coloc R package for Bayesian colocalization analysis^60^. mQTL summary statistics were tested for each of the CpG sites in the *SNCA* region separately, overlapping to loci present in both mQTL results and GWAS summary statistics tested. The coloc analysis was run using the full effect estimates, variance estimates, minor allele frequency and N values for each summary statistic set. An H4 prior probability of > 0.9 was interpreted as evidence of colocalization. Given the *SNCA* genetic risk profile from the largest GWAS of PD to date represent multiple nominated variants, we utilized summary statistics of a conditioned analysis of the region to test the independent effects of the primary and secondary genetic association signals in PD^61^. For this analysis, the results conditioned on the strongest GWAS locus rs356182 were labelled the “PD 5’ Signal”, whilst the results conditioned on the secondary locus rs763443 were labelled the “PD 3’ Signal”. The third independent signal at rs280004 and the unconditioned summary statistics were also tested. For LBD, the largest GWAS to date was utilized^5^.

### SNCA gene expression analysis

To analyze the effect of DNA methylation and genotype at the nominated *SNCA* locus with gene expression, short- and long-read RNA sequencing data from previously published PFC samples were sourced. For the short read expression analysis, data from the ROSMAP cohort was sourced as outlined in Laroche et al 2025 and covered matched Illumina 450K methylation array data, bulk short-read RNA sequencing and imputed genotype data for 211 samples^19^. Transcripts per Million (TPM) normalized expression for *SNCA* and *SNCA-AS1* were extracted and tested for association against DNA methylation at cg01966878 and genotype at rs7680557 (coded as number of C-alleles) using linear regression analysis, controlling for covariates of age, sex, PMI, RNA integrity number (RIN) and DNA methylation derived estimates for the five cell proportions.

For assessment of *SNCA* transcript isoforms, 49 previously published data from PFC of the BDR cohort with targeted Oxford Nanopore long-read RNA sequencing was sourced^20^. For this analysis, only transcripts with prior annotation were included and were required to fulfil a full-splicing match criteria in SQANTI3 for inclusion. Where multiple isoforms for an individual annotated transcript were present, these were concatenated into a single isoform annotation. Transcript isoforms were counts per million normalized (CPM) using the cpm() function in edgeR. Each called transcript isoform of *SNCA* was tested for its association with DNA methylation at cg01966878 and genotype at rs7680557 using the same covariates and methods as in the ROSMAP short-read RNA sequencing analysis.

## Data Availability

Methylation data, genotyping data and available phenotypic data used in primary analyses for the UK Brain Bank Network cohort will be made available on the Gene Expression Omnibus (GEO) Platform upon peer review and final publication. Data for the Netherlands Brain Bank cohort is available via GEO at accession number GSE203332. Data for the Brains for Dementia Research cohort is available via GEO at accession number GSE197305 and additional donor data is available via Dementia Platform UK (https://portal.dementiasplatform.uk/).
Genome-wide summary statistics for meta-analyses performed in this study are available at https://github.com/JoshHarveyGit/LB_Meta_Analysis.

https://github.com/JoshHarveyGit/LB_Meta_Analysis

## DATA AVAILABILITY

Methylation data, genotyping data and available phenotypic data used in primary analyses for the UKBBN cohort will be made available on the Gene Expression Omnibus (GEO) Platform upon final publication. Data for the NBB cohort is available via GEO at accession number GSE203332. Data for the BDR cohort is available via GEO at accession number GSE197305 and additional donor data is available via Dementia Platform UK (https://portal.dementiasplatform.uk/).

Genome-wide summary statistics for meta-analyses performed in this study are available at https://github.com/UoE-Dementia-Genomics/LB-Meta-Analysis

## CODE AVAILABILITY

Analysis scripts used in this manuscript are available at https://github.com/UoE-Dementia-Genomics/LB-Meta-Analysis

## ACKNOWLEDGEMENTS

We acknowledge the brain banks used in the sourcing of tissue, clinical and neuropathological data used in this study. These include the Oxford, UCL Queen’s Square, Imperial College London Parkinson’s disease Brain Bank and Newcastle Brain Banks and we thank the donors who contributed to this study.

## AUTHOR INFORMATION

JH, JI and ARS conducted laboratory experiments generating the DNA methylation data. JH undertook data analysis and bioinformatics, with support from MK, BC, RGS, SKL, VL and GW. ZJ, EDP-F, TW, DL, DG, HB, JA, WDJVB and AT supported with sample characterization and selection. GS, ED, LP and JM provided data for the meta-analysis. LC, CB, JO’B, DA and KL conceived the project. EP and KL supervised the project. JH, JI, EP and KL drafted the manuscript. All authors read and approved the final submission.

## ETHICS DECLARATIONS

The authors declare no competing interests

## REFERENCES

1. (!!! INVALID CITATION !!! 1-3).

2. McKeith, I.G. et al. Diagnosis and management of dementia with Lewy bodies: Fourth consensus report of the DLB Consortium. Neurology 89, 88–100 (2017).

3. Jellinger, K.A. & Korczyn, A.D. Are dementia with Lewy bodies and Parkinson’s disease dementia the same disease? BMC Med 16, 34 (2018).

4. Kim, J.J. et al. Multi-ancestry genome-wide association meta-analysis of Parkinson’s disease. Nat Genet 56, 27–36 (2024).

5. Chia, R. et al. Genome sequencing analysis identifies new loci associated with Lewy body dementia and provides insights into its genetic architecture. Nat Genet 53, 294–303 (2021).

6. Tsalenchuk, M., Gentleman, S.M. & Marzi, S.J. Linking environmental risk factors with epigenetic mechanisms in Parkinson’s disease. NPJ Parkinsons Dis 9, 123 (2023).

7. Ben-Shlomo, Y. et al. The epidemiology of Parkinson’s disease. Lancet 403, 283–292 (2024).

8. An, D. & Xu, Y. Environmental risk factors provoke new thinking for prevention and treatment of dementia with Lewy bodies. Heliyon 10, e30175 (2024).

9. Lunnon, K. et al. Methylomic profiling implicates cortical deregulation of ANK1 in Alzheimer’s disease. Nat Neurosci 17, 1164–70 (2014).

10. De Jager, P.L. et al. Alzheimer’s disease: early alterations in brain DNA methylation at ANK1, BIN1, RHBDF2 and other loci. Nat Neurosci 17, 1156–63 (2014).

11. Smith, A.R. et al. Parallel profiling of DNA methylation and hydroxymethylation highlights neuropathology-associated epigenetic variation in Alzheimer’s disease. Clin Epigenetics 11, 52 (2019).

12. Smith, R.G. et al. A meta-analysis of epigenome-wide association studies in Alzheimer’s disease highlights novel differentially methylated loci across cortex. Nat Commun 12, 3517 (2021).

13. Pihlstrom, L. et al. Epigenome-wide association study of human frontal cortex identifies differential methylation in Lewy body pathology. Nat Commun 13, 4932 (2022).

14. Shireby, G. et al. DNA methylation signatures of Alzheimer’s disease neuropathology in the cortex are primarily driven by variation in non-neuronal cell-types. Nat Commun 13, 5620 (2022).

15. Robinson, J.L. et al. Neurodegenerative disease concomitant proteinopathies are prevalent, age-related and APOE4-associated. Brain 141, 2181–2193 (2018).

16. Wheildon, G. et al. DNA methylation profiling in Huntington’s disease reveals disease associated changes in the striatum. (2025).

17. Bellenguez, C. et al. New insights into the genetic etiology of Alzheimer’s disease and related dementias. Nat Genet 54, 412–436 (2022).

18. Kunkle, B.W. et al. Genetic meta-analysis of diagnosed Alzheimer’s disease identifies new risk loci and implicates Abeta, tau, immunity and lipid processing. Nat Genet 51, 414–430 (2019).

19. Laroche, V.T. et al. Epigenomic subtypes of late-onset Alzheimer’s disease reveal distinct microglial signatures. bioRxiv, 2025.03.15.643144 (2025).

20. Leung, S.K. et al. Long-read transcript sequencing identifies differential isoform expression in the entorhinal cortex in a transgenic model of tau pathology. Nat Commun 15, 6458 (2024).

21. Vukojevic, K., Soljic, V., Martinovic, V., Raguz, F. & Filipovic, N. The Ubiquitin-Associated and SH3 Domain-Containing Proteins (UBASH3) Family in Mammalian Development and Immune Response. Int J Mol Sci 25(2024).

22. Zhu, B. et al. Single-cell transcriptomic and proteomic analysis of Parkinson’s disease brains. Sci Transl Med 16, eabo1997 (2024).

23. Feleke, R. et al. Cross-platform transcriptional profiling identifies common and distinct molecular pathologies in Lewy body diseases. Acta Neuropathol 142, 449–474 (2021).

24. Sarnowski, C. et al. Meta-analysis of genome-wide association studies identifies ancestry-specific associations underlying circulating total tau levels. Commun Biol 5, 336 (2022).

25. Liu, J. et al. Platelet Activating Factor Receptor Exaggerates Microglia-Mediated Microenvironment by IL10-STAT3 Signaling: A Novel Potential Biomarker and Target for Diagnosis and Treatment of Alzheimer’s Disease. Front Aging Neurosci 14, 856628 (2022).

26. Hans, S. et al. Polar lipids modify Alzheimer’s Disease pathology by reducing astrocyte pro-inflammatory signaling through platelet-activating factor receptor (PTAFR) modulation. Lipids Health Dis 23, 113 (2024).

27. Duggan, M.R. et al. Proteome-wide analysis identifies plasma immune regulators of amyloid-beta progression. Brain Behav Immun 120, 604–619 (2024).

28. Repudi, S. et al. Neuronal deletion of Wwox, associated with WOREE syndrome, causes epilepsy and myelin defects. Brain 144, 3061–3077 (2021).

29. Liu, G. et al. Genome-wide survival study identifies a novel synaptic locus and polygenic score for cognitive progression in Parkinson’s disease. Nat Genet 53, 787–793 (2021).

30. Gustavsson, E.K. et al. RAB32 Ser71Arg in autosomal dominant Parkinson’s disease: linkage, association, and functional analyses. Lancet Neurol 23, 603–614 (2024).

31. Ai, S.X. et al. Hypomethylation of SNCA in blood of patients with sporadic Parkinson’s disease. J Neurol Sci 337, 123–8 (2014).

32. Jowaed, A., Schmitt, I., Kaut, O. & Wullner, U. Methylation regulates alpha-synuclein expression and is decreased in Parkinson’s disease patients’ brains. J Neurosci 30, 6355–9 (2010).

33. Matsumoto, L. et al. CpG demethylation enhances alpha-synuclein expression and affects the pathogenesis of Parkinson’s disease. PLoS One 5, e15522 (2010).

34. Desplats, P. et al. Alpha-synuclein sequesters Dnmt1 from the nucleus: a novel mechanism for epigenetic alterations in Lewy body diseases. J Biol Chem 286, 9031–7 (2011).

35. Kantor, B. et al. Downregulation of SNCA Expression by Targeted Editing of DNA Methylation: A Potential Strategy for Precision Therapy in PD. Mol Ther 26, 2638–2649 (2018).

36. Pihlstrom, L., Berge, V., Rengmark, A. & Toft, M. Parkinson’s disease correlates with promoter methylation in the alpha-synuclein gene. Mov Disord 30, 577–80 (2015).

37. Krohn, L. et al. Genome-wide association study of REM sleep behavior disorder identifies polygenic risk and brain expression effects. Nat Commun 13, 7496 (2022).

38. Alvarez Jerez, P., et al. Characterizing a complex CT-rich haplotype in intron 4 of SNCA using large-scale targeted amplicon long-read sequencing. NPJ Parkinsons Dis 10, 136 (2024).

39. Gu, J. et al. Cell-Type Specific Changes in DNA Methylation of SNCA Intron 1 in Synucleinopathy Brains. Front Neurosci 15, 652226 (2021).

40. Wainberg, M., Andrews, S.J. & Tripathy, S.J. Shared genetic risk loci between Alzheimer’s disease and related dementias, Parkinson’s disease, and amyotrophic lateral sclerosis. Alzheimers Res Ther 15, 113 (2023).

41. Stolp Andersen, M., et al. Dissecting the limited genetic overlap of Parkinson’s and Alzheimer’s disease. Ann Clin Transl Neurol 9, 1289–1295 (2022).

42. Olney, K.C. et al. Distinct transcriptional alterations distinguish Lewy body disease from Alzheimer’s disease. Brain 148, 69–88 (2025).

43. Attems, J. et al. Neuropathological consensus criteria for the evaluation of Lewy pathology in post-mortem brains: a multi-centre study. Acta Neuropathol 141, 159–172 (2021).

44. Müller, J. et al. A Cell Type Enrichment Analysis Tool for Brain DNA Methylation Data (CEAM). bioRxiv, 2025.07.08.663671 (2025).

45. Beach, T.G. et al. Unified staging system for Lewy body disorders: correlation with nigrostriatal degeneration, cognitive impairment and motor dysfunction. Acta Neuropathol 117, 613–34 (2009).

46. Alafuzoff, I. et al. Staging/typing of Lewy body related alpha-synuclein pathology: a study of the BrainNet Europe Consortium. Acta Neuropathol 117, 635–52 (2009).

47. Aryee, M.J. et al. Minfi: a flexible and comprehensive Bioconductor package for the analysis of Infinium DNA methylation microarrays. Bioinformatics 30, 1363–9 (2014).

48. Pidsley, R. et al. A data-driven approach to preprocessing Illumina 450K methylation array data. BMC Genomics 14, 293 (2013).

49. Vellame, D.S. et al. Uncertainty quantification of reference-based cellular deconvolution algorithms. Epigenetics 18, 2137659 (2023).

50. Hannon, E. et al. Quantifying the proportion of different cell types in the human cortex using DNA methylation profiles. BMC Biol 22, 17 (2024).

51. Harvey, J. et al. Machine learning-based prediction of cognitive outcomes in de novo Parkinson’s disease. NPJ Parkinsons Dis 8, 150 (2022).

52. Balduzzi, S., Rucker, G. & Schwarzer, G. How to perform a meta-analysis with R: a practical tutorial. Evid Based Ment Health 22, 153–160 (2019).

53. Fodder, K. et al. Brain DNA methylomic analysis of frontotemporal lobar degeneration reveals OTUD4 in shared dysregulated signatures across pathological subtypes. Acta Neuropathol 146, 77–95 (2023).

54. Mansell, G. et al. Guidance for DNA methylation studies: statistical insights from the Illumina EPIC array. BMC Genomics 20, 366 (2019).

55. Pedersen, B.S., Schwartz, D.A., Yang, I.V. & Kechris, K.J. Comb-p: software for combining, analyzing, grouping and correcting spatially correlated P-values. Bioinformatics 28, 2986–8 (2012).

56. Ren, X. & Kuan, P.F. methylGSA: a Bioconductor package and Shiny app for DNA methylation data length bias adjustment in gene set testing. Bioinformatics 35, 1958–1959 (2019).

57. Sayols, S. rrvgo: a Bioconductor package for interpreting lists of Gene Ontology terms. MicroPubl Biol 2023(2023).

58. Shabalin, A.A. Matrix eQTL: ultra fast eQTL analysis via large matrix operations. Bioinformatics 28, 1353–8 (2012).

59. Poole, W., Gibbs, D.L., Shmulevich, I., Bernard, B. & Knijnenburg, T.A. Combining dependent P-values with an empirical adaptation of Brown’s method. Bioinformatics 32, i430–i436 (2016).

60. Wallace, C. Statistical testing of shared genetic control for potentially related traits. Genet Epidemiol 37, 802–13 (2013).

61. Pihlstrom, L. et al. A comprehensive analysis of SNCA-related genetic risk in sporadic parkinson disease. Ann Neurol 84, 117–129 (2018).

