## Supplementary Figures for "Interrogating DNA methylation associated with Lewy body pathology in a cross brain-region and multi-cohort study"

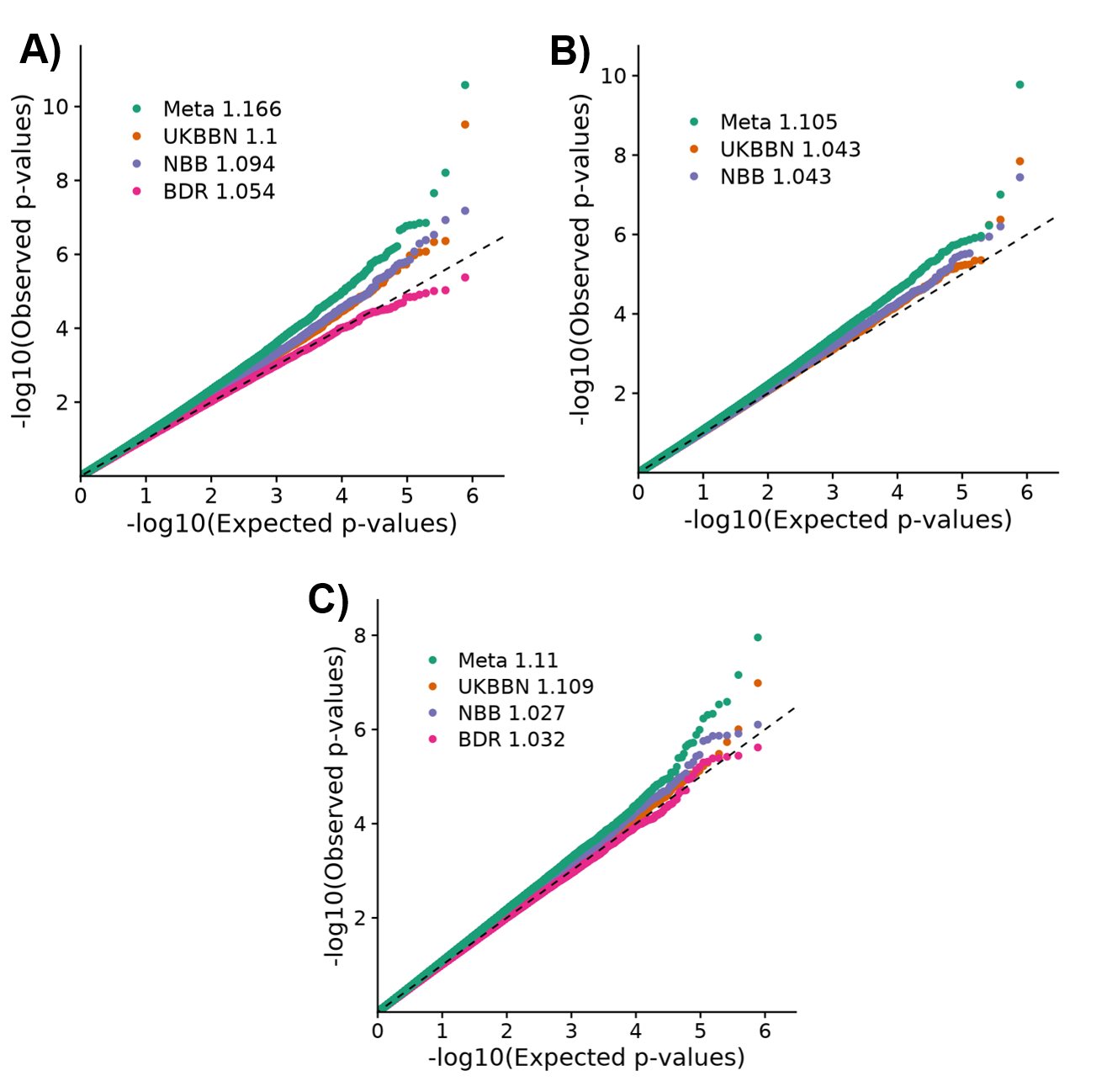

**Supplementary Figure 1.** **Quantile-Quantile (QQ) plots showing genomic inflation for each cohort and final meta-analysis fixed effect p-values.** Shown are **A)** primary full cohort LB pathology meta-analysis, **B)** secondary pure LB pathology subset cohort meta-analysis and **C)** Thal-controlled LB pathology meta-analysis. Abbreviations: Meta = Meta analysis fixed effect p-values, UKBBN = UK Brain Bank Network mixed effect model p-values, NBB = Netherlands Brain Bank linear model p-values, BDR = Brains for Dementia Research linear model p-values. Shown next to each cohort and test is the Lambda genomic inflation value for each EWAS p-value set tested.

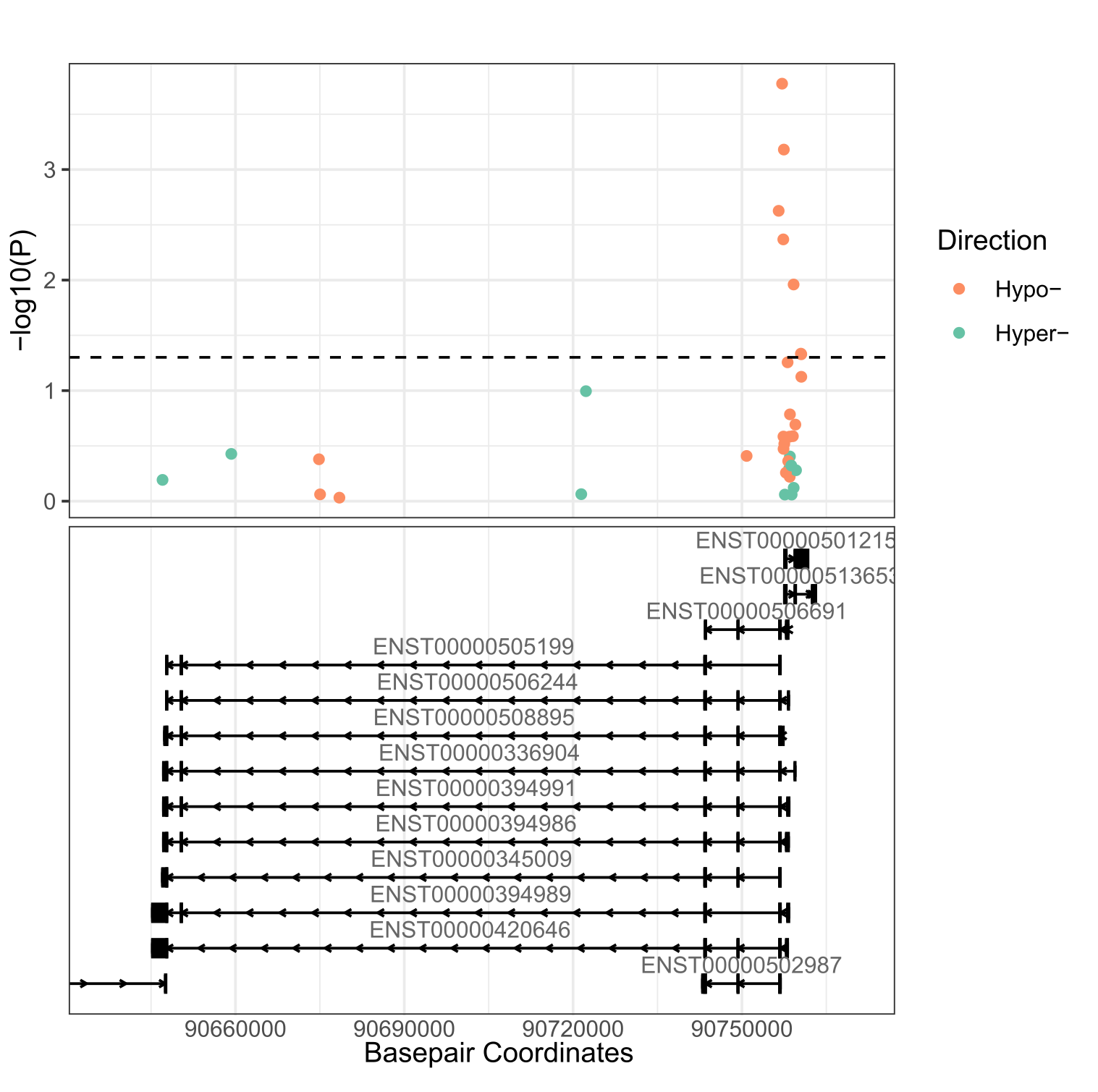
**Supplementary Figure 2.** **Seven probes in the *SNCA* gene showed nominally significant hypomethylation in the primary full cohort LB pathology meta-analysis.** Robust rank aggregation (RRA) of the meta-analysis results highlighted 245 significant genes, including *SNCA*. Shown are all probes annotated to the *SNCA* gene region (Chr4:90637041-90770556), with those showing hypomethylation in association with LB Braak stage colored in orange, and those showing hypermethylation colored in green. The black dashed horizontal line denotes nominal significance (p < 0.05). Underneath the *SNCA* and *SNCA-AS1* gene transcripts are annotated.

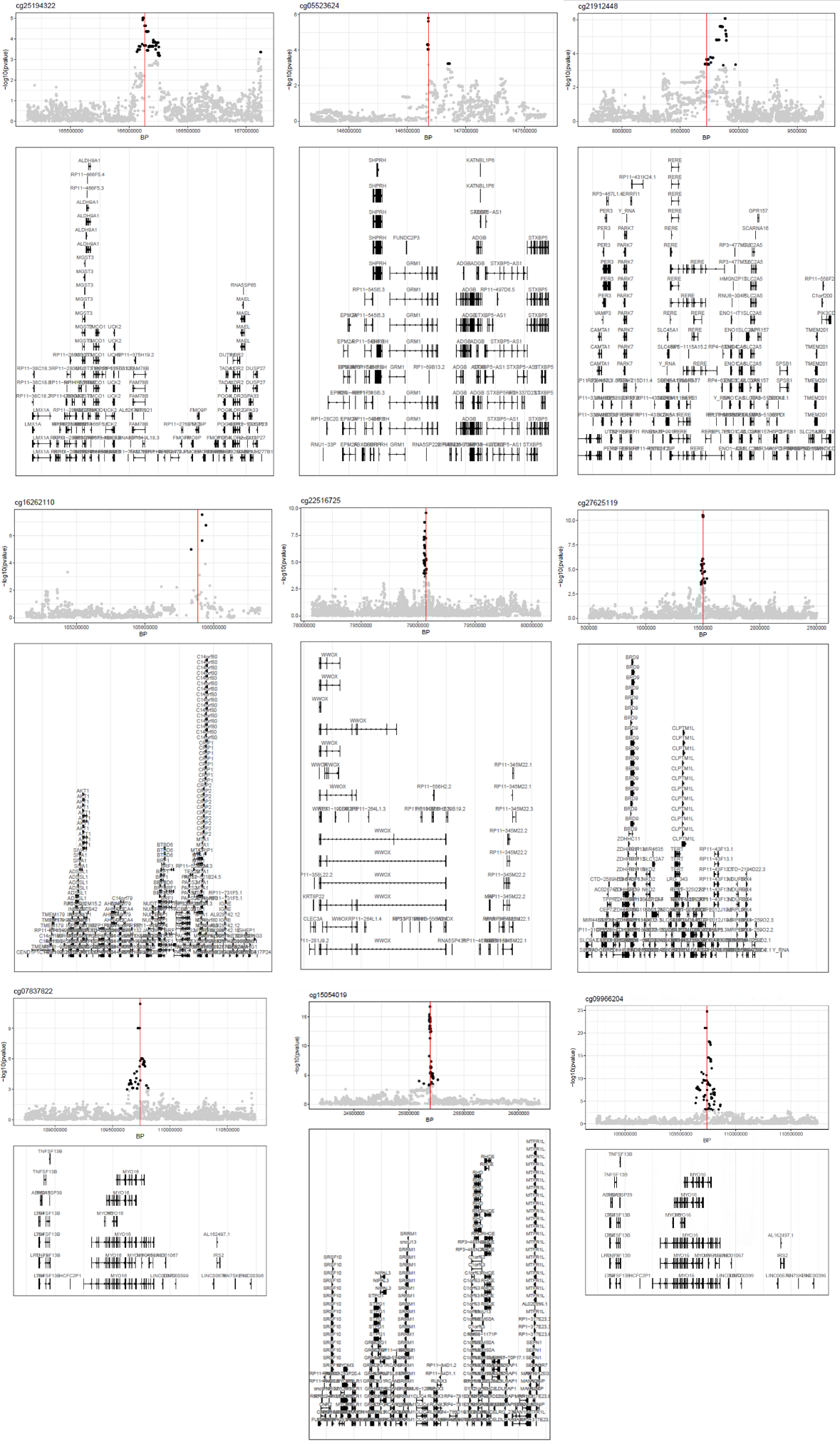

**Supplementary Figure 3. Significant *cis*-methylation quantitative trait loci (mQTL) relationships in the UKBBN cohort for FDR significant loci identified in the primary full cohort LB pathology meta-analysis.** X-axis shows base-pair coordinate, with y-axis showing unadjusted -log10(p-value) for genotype-methylation associations. Red vertical bar shows location of the methylation site. For each mini-manhattan plot, tracks showing genomic context is shown. Dark points represent FDR-significant mQTL associations with grey points showing non-significant associations.

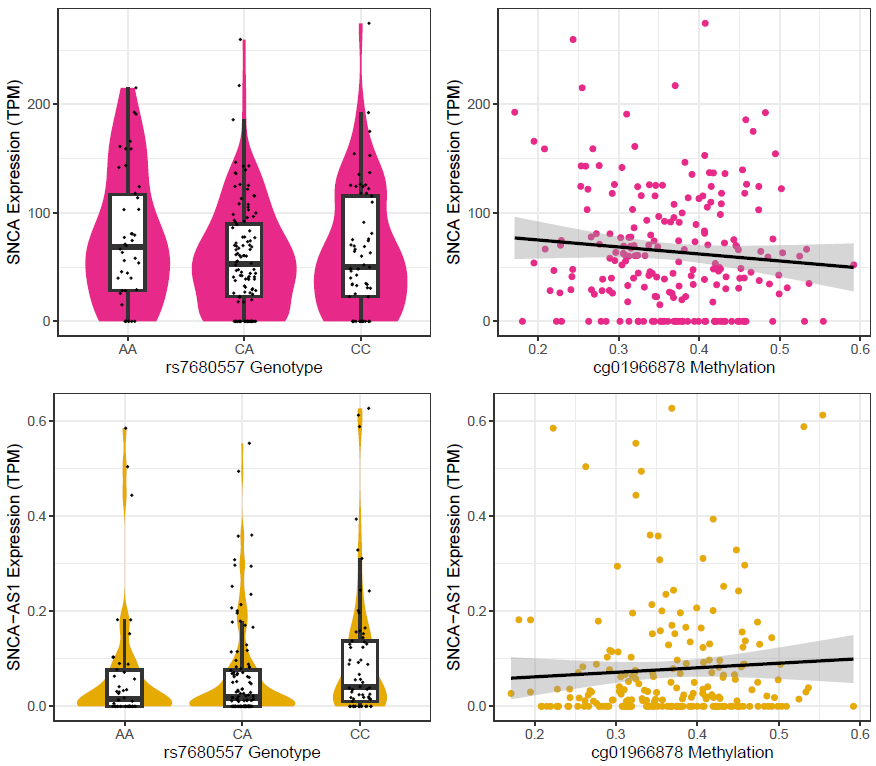

**Supplementary Figure 4**. **Exploration of *SNCA* and *SNCA-AS1* transcripts using short read RNA-sequencing data in the ROSMAP cohort.** Shown are transcript-per-million (TPM) normalized expression against allele for the lead DLB associated SNP rs7680557 for **A)** *SNCA* and **B)** *SNCA-AS1*. Scatter plots are shown for **C)** *SNCA* and **D)** *SNCA-AS1* gene expression against DNA-methylation at cg019668787 within the *SNCA* gene region. The line shows the linear model-based line of best fit.
